# Feasibility Study of Ferumoxtyol for Contrast-enhanced MRI of Retroplacental Clear Space Disruption in Placenta Accreta Spectrum (PAS)

**DOI:** 10.1101/2023.03.20.23287436

**Authors:** Andrew A. Badachhape, Brian Burnett, Prajwal Bhandari, Laxman Devkota, Rohan Bhavane, Ketan B. Ghaghada, Chandrasekhar Yallampalli, Karin A. Fox, Ananth V. Annapragada

## Abstract

**Introduction:** Placenta accreta spectrum (PAS) occurs when the placenta is pathologically adherent to the myometrium. An intact retroplacental clear space (RPCS) is a marker of normal placentation, but visualization with conventional imaging techniques is a challenge. In this study, we investigate use of an FDA-approved iron oxide nanoparticle, ferumoxytol, for contrast-enhanced magnetic resonance imaging of the RPCS in mouse models of normal pregnancy and PAS. We then demonstrate the translational potential of this technique in human patients presenting with severe PAS (FIGO Grade 3C), moderate PAS (FIGO Grade 1), and no PAS.

**Methods:** A T1-weighted gradient recalled echo (GRE) sequence was used to determine the optimal dose of ferumoxytol in pregnant mice. Pregnant Gab3^-/-^ mice, which demonstrate placental invasion, were then imaged at day 16 of gestation alongside wild-type (WT) pregnant mice which do not demonstrate invasion. Signal-to-noise ratio (SNR) was computed for placenta and RPCS for all fetoplacental units (FPUs) with ferumoxytol-enhanced magnetic resonance imaging (Fe-MRI) and used for the determination of contrast-to-noise ratio (CNR). Fe-MRI was also performed in 3 pregnant subjects using standard T1 and T2 weighted sequences and a 3D magnetic resonance angiography (MRA) sequence. RPCS volume and relative signal were calculated in all three subjects.

**Results:** Ferumoxytol administered at 5 mg/kg produced strong T1 shortening in blood and led to strong placental enhancement in Fe-MRI images. Gab3^-/-^ mice demonstrated loss of hypointense region characteristic of the RPCS relative to WT mice in T1w Fe-MRI. CNR between RPCS and placenta was lower in FPUs of Gab3^-/-^ mice compared to WT mice, indicating higher degrees of vascularization and interruptions throughout the space. In human patients, Fe-MRI at a dose of 5 mg/kg enabled high uteroplacental vasculature signal and quantification of the volume and signal profile in severe and moderate invasion of the placenta relative to a non-PAS case.

**Discussion:** Ferumoxytol, an FDA-approved iron oxide nanoparticle formulation, enabled visualization of abnormal vascularization and loss of uteroplacental interface in a murine model of PAS. The potential of this non-invasive visualization technique was then further demonstrated in human subjects. Diagnosis of placental invasion using Fe-MRI may provide a sensitive method for clinical detection of PAS.

## Introduction

Placenta Accreta Spectrum (PAS), where the placenta becomes pathologically adherent to the uterine myometrium and complicates delivery at term, has increased in incidence [1–3] and remains a difficult condition to assess antenatally [4–7]. One specific concern is related to conventional magnetic resonance imaging (MRI), which often follows an unclear ultrasound read; however, this practice is unreliable and may lead to unwarranted changes in clinical management [8]. Non-contrast MRI relies on indirect signs of pathology in the setting of PAS, specifically dark intraplacental bands, confirmation of placenta previa, and large changes in placental tissue architecture [9].

There remains a critical need for a non-invasive method to assess the interface between the uterus and placenta, often termed the junctional zone, or retroplacental clear space [10,11]. Clear visualization of this space may provide a more accurate method for diagnosing PAS disorders and quantifying the severity of the condition. Previous work by our group has explored novel liposomal-based contrast agents for visualizing the RPCS in rodent models of pregnancy and PAS [12–15]. While these agents have strong T1 relaxive properties and spare the fetus from gadolinium exposure [13,14], they are not FDA approved, and substituting them with an FDA-approved agent that provides similar T1-relaxive properties could have immediate clinical applications. One potential substitute is ferumoxytol, a superparamagnetic iron oxide nanoparticle agent approved for the treatment of iron-deficiency anemia, including in pregnant patients [16]. Patients with PAS are at increased risk for intrapartum hemorrhage and intravenous iron may be considered during antenatal optimization of hemoglobin. Ferumoxytol has been used off-label as a contrast agent for vascular imaging, perfusion imaging, and other applications[17]. Multiple sites have begun studies to determine the efficacy of this technique for diagnosing PAS and other placental disorders [18–20].

In this paper, we seek to assess the feasibility of ferumoxytol-enhanced magnetic resonance imaging (Fe-MRI) for the visualization RPCS disruption in PAS. Building upon our previous observation of strong placental signal increase following administration of liposomal-Gd agents, we hypothesize that we can achieve similar performance with ferumoxytol and visualize the RPCS in both preclinical and clinical experiments. In this work, we begin with preclinical studies in rodent models of pregnancy and PAS to optimize dose and assess quantitative metrics that can identify and grade PAS disorders. We then translate this work into three clinical cases of PAS: (1) FIGO Grade 3C, (2) FIGO Grade 1, and (3) absent [21–23].

## Methods

### Animal Model and Breeding Study

The Institutional Animal Care and Use Committee at Baylor College of Medicine approved all animal studies. Both C57BL/6 wild type (WT) and Gab3^-/-^ mice were used in this study. Gab3^-/-^ mice were bred in-house from a breed pair generously provided by Dr. Helen Jones at the University of Cincinnati [15,24]. A total of 9 WT mice and 3 Gab3^-/-^ mice were used. All mice were 8-12 weeks old and approximately ∼20-30g body weight before pregnancy. All Gab3^-/-^ female mice were mated with Gab3^-/-^ male mice and all WT female mice were mated with WT male mice. The first day of gestation, designated as 0.5, was when a vaginal copulation plug was detected. Fe-MR imaging was performed on day 12.5 of pregnancy (E12.5) in 3 WT mice. A separate dose-ranging study was performed at day 16.5 of pregnancy (E16.5) in 3 WT mice. Finally, a comparative imaging study was performed in 3 WT and 3 Gab3^-/-^ mice at E16.5. A total of 29 FPUs were examined in WT mice at E12.5. A total of 18 Gab3^-/-^ FPUs and 24 WT FPUs were examined and compared at E16.5.

### Magnetic Resonance Imaging (MRI) - Mice

Mouse imaging was performed on a 1T permanent MRI scanner (M2 system, Aspect Imaging, Shoham, Israel) with a 35 mm transmit-receive RF volume coil, following methods described previously [12,14,25]. Briefly, mice were sedated using 3% isoflurane, placed on the MRI animal bed, and then maintained at 1-2% isoflurane delivered using a nose cone setup. Breathing rate was monitored through pressure pad placed below the abdomen. Ferumoxytol was intravenously administered to pregnant mice in one of three doses (1 mg/kg, 5 mg/kg, or 10 mg/kg) via tail vein in the dose ranging study. Ferumoxytol was administered at a dose of 5 mg/kg in all other studies.

All animals underwent pre-contrast and post-contrast scans. T1-weighted (T1w) scans were acquired with a 3D gradient echo (GRE) sequence. The 3D GRE sequence had the following parameters: TR = 20 ms, TE = 3.5 ms, slice thickness = 0.3 mm, in-plane resolution = 300 μm x 300 μm, scan time ∼ 6 min. The same T1w-GRE sequences were used to acquire post-contrast scans.

Three acquisitions were performed for the 3D GRE scan protocol. Magnitude-sum averages were calculated in Matlab® (version R2015a, MathWorks©, Natick, MA) using acquisitions that showed little to no motion artifact.

### Ultrasound and MR Imaging – Human

The Institutional Review Board of Baylor College of Medicine gave ethical approval for this work. Patients who had been prescribed intravenous iron supplementation, and at risk for PAS by patient history and ultrasound findings, per ACOG guidelines [26,27], were recruited for this study. Patients with a partially filled bladder underwent an abdominal ultrasound examination using a GE-10 ultrasound scanner. Acquisition included full sweep 2-D greyscale and color Doppler images in the parasagittal and transverse planes. Transverse sweeps of the parametrial interface were performed. Transvaginal imaging and 3D rendering were performed as necessary. A parasagittal view of the maternal pelvis was evaluated to observe intra-placental vasculature and the serosa-bladder interface.

MRI was performed on a 1.5T Phillips Ingenia scanner. Patients underwent 1)T2-weighted turbo spin echo with the following parameters: TR: 800 ms, TE: 80 ms, slice thickness: 5 mm, in-plane resolution: 0.8 mm x 0.8 mm, scan time ∼1 min; 2) mDixon sequence with the following parameters: TR: 5.6 ms, TE = 1.75 ms, slice thickness = 5 mm, in-plane resolution: 0.8 mm x 0.8 mm, scan time ∼1.5 min; and 3)3D magnetic resonance angiography with the following parameters: TR: 5.2 ms, TE: 1.4 ms, slice thickness = 0.6 mm, in-plane resolution: 0.6 mm x 0.6 mm, scan time ∼3 min. Following baseline, pre-contrast scans, subjects were intravenously administered ferumoxytol (5 mg/kg) as a slow infusion and then underwent post-contrast scans using the same scan protocol as the pre-contrast scans.

### Image Analysis

All MR images were analyzed with OsiriX (version 5.8.5, 64-bit) and MATLAB (version 2015a). In mice, regions-of-interest were manually drawn to segment the placenta (P) and the retroplacental clear space (RPCS) in GRE images. Volumetric and signal data from these ROIs were determined for the segmented structures, and the data is presented in terms of average and standard deviation. Similar ROIs were drawn in clinical data using the 3D MRA image set to evaluate placental-RPCS CNR and volume. All statistical analysis was performed using a Wilcoxon rank-sum test.

## Results

Dose-ranging studies in healthy C57BL/6 pregnant mice demonstrate that a moderate dose of 5 mg/kg results in superior T1 relaxation at a 1T field strength (**Figure 1**). At doses greater than 5 mg/kg, strong T2 effect of ferumoxytol overrides T1 relaxive effects of nanoparticle, resulting in lower signal. In vivo studies in pregnant C57BL/6 mice using 5 mg/kg dose of ferumoxytol demonstrated high signal-to-noise ratio (SNR) in the placenta, which enabled visualization of the RPCS at E12.5, the earliest timepoint that the RPCS is visible, and at E16.5, where RPCS volume reaches a maximum [12] (**Figure 2**). After demonstrating the feasibility of T1-weighted Fe-MRI, we performed a comparative study of the Gab3^-/-^ mouse model with C57BL/6 WT controls where both groups of animals were administered 5 mg/kg of ferumoxytol to visualize the RPCS. In the Gab3^-/-^ model, frank disruption of the RPCS was visible in Fe-MR images, and quantification of the space revealed heterogeneous signal profile in the RPCS (**Figure 3**). These results indicate disruption of hypointense RPCS region, which led to a lower conspicuity score (Placenta-RPCS CNR) in the Gab3^-/-^ model relative to WT controls.

**Figure 1:**
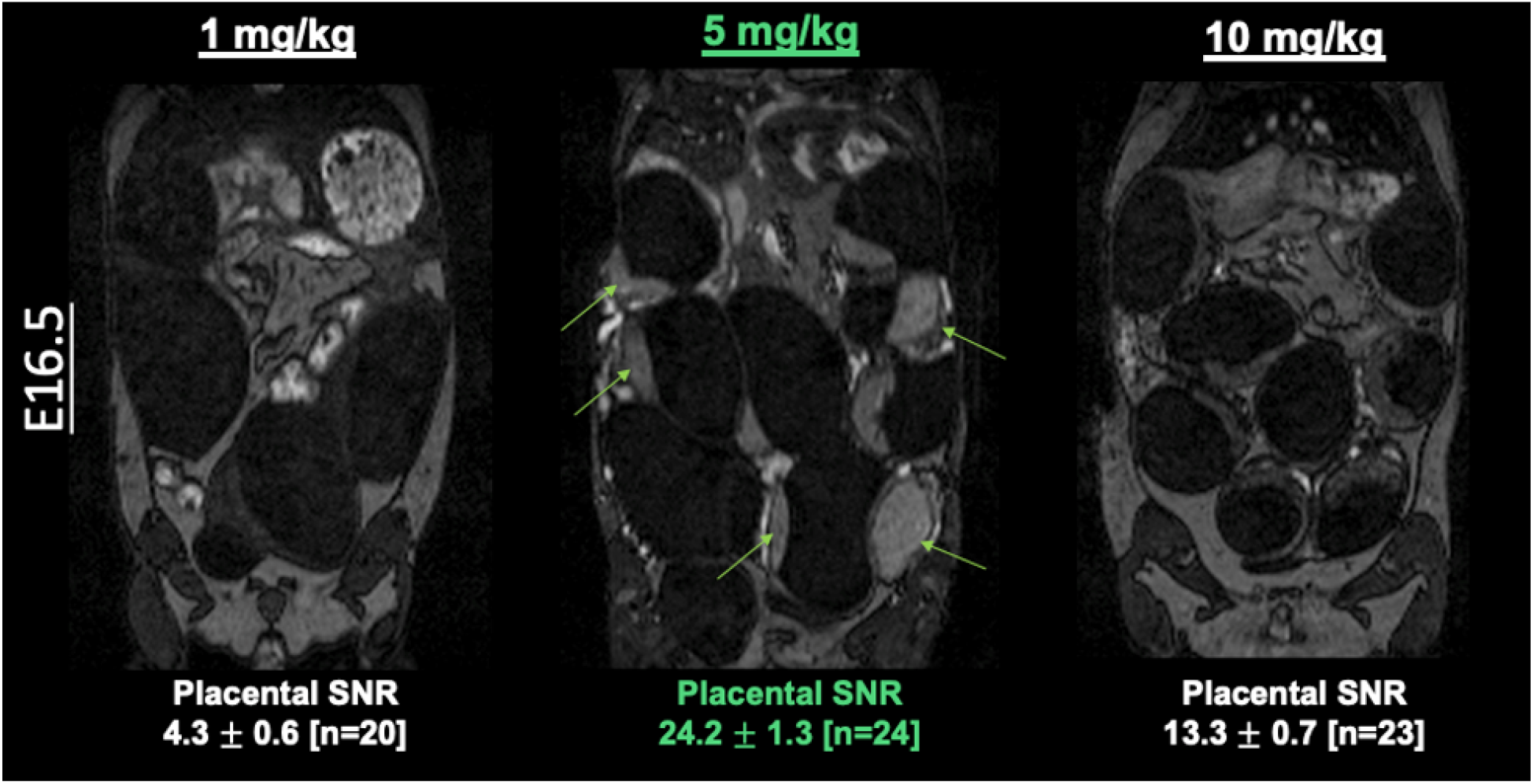
T1-weighted gradient recalled echo (GRE) images at E16.5 demonstrate that a moderate ferumoxytol dose of 5 mg/kg provides optimal T1-weighting. Placental signal-to-noise ratio (SNR) was higher at 5 mg/kg than at 1 mg/kg or 10 mg/kg.

**Figure 2:**
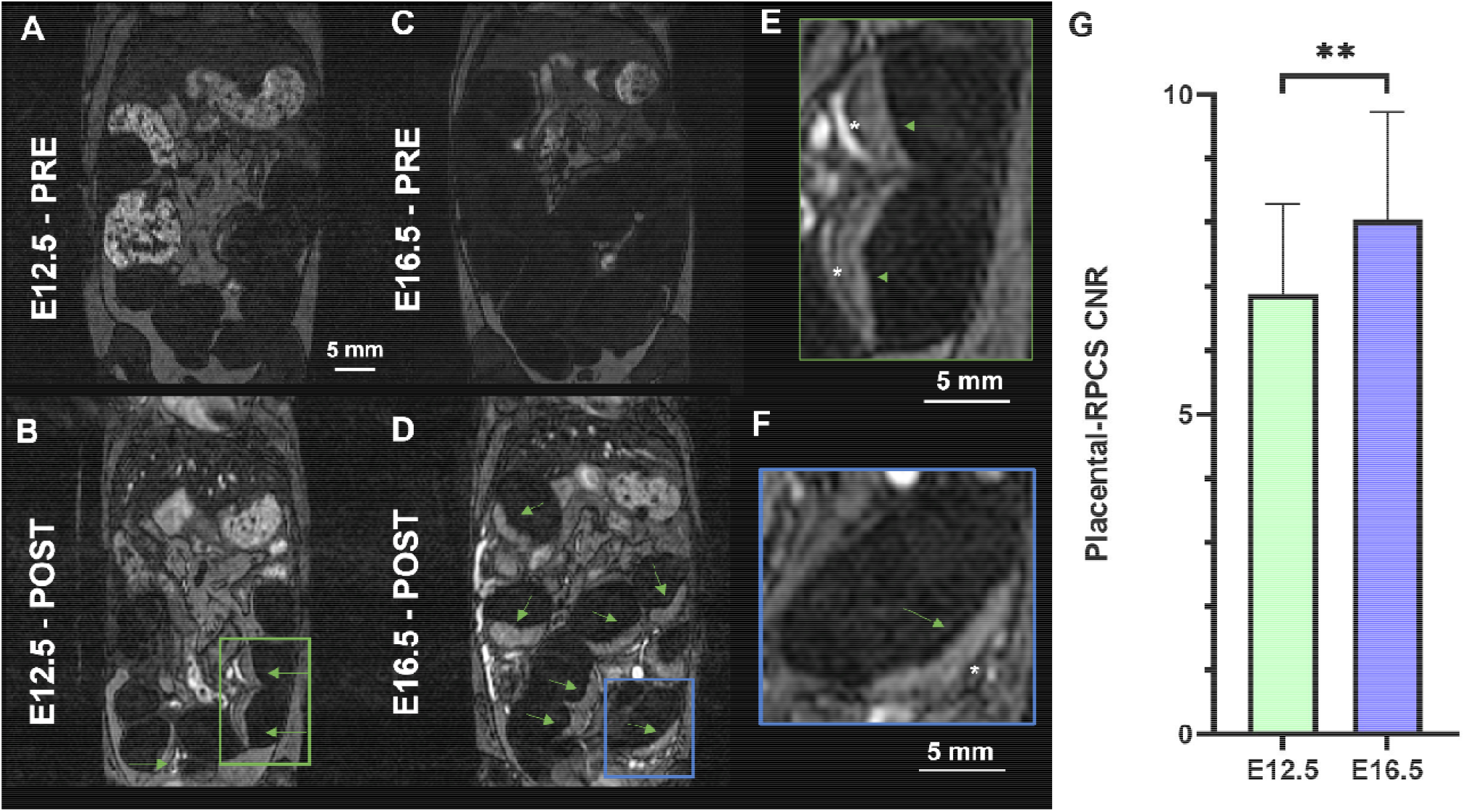
Ferumoxytol-enhanced MRI enables visualization of the retroplacental clear space (RPCS). (**A, C**) Pre-contrast T1-weighted gradient recalled echo (GRE) images acquired prior to ferumoxytol administration at E12.5 and E16.5 demonstrate little to no signal in the placenta (arrows) relative to (**B, D**) post-contrast images at E12.5 and E16.5. (**E, F**) Higher magnification images demonstrate the RPCS (*). (**G**) The contrast-to-noise ratio of the RPCS relative to the placenta is significantly higher (p<0.005) at E16.5 relative to E12.5.

**Figure 3:**
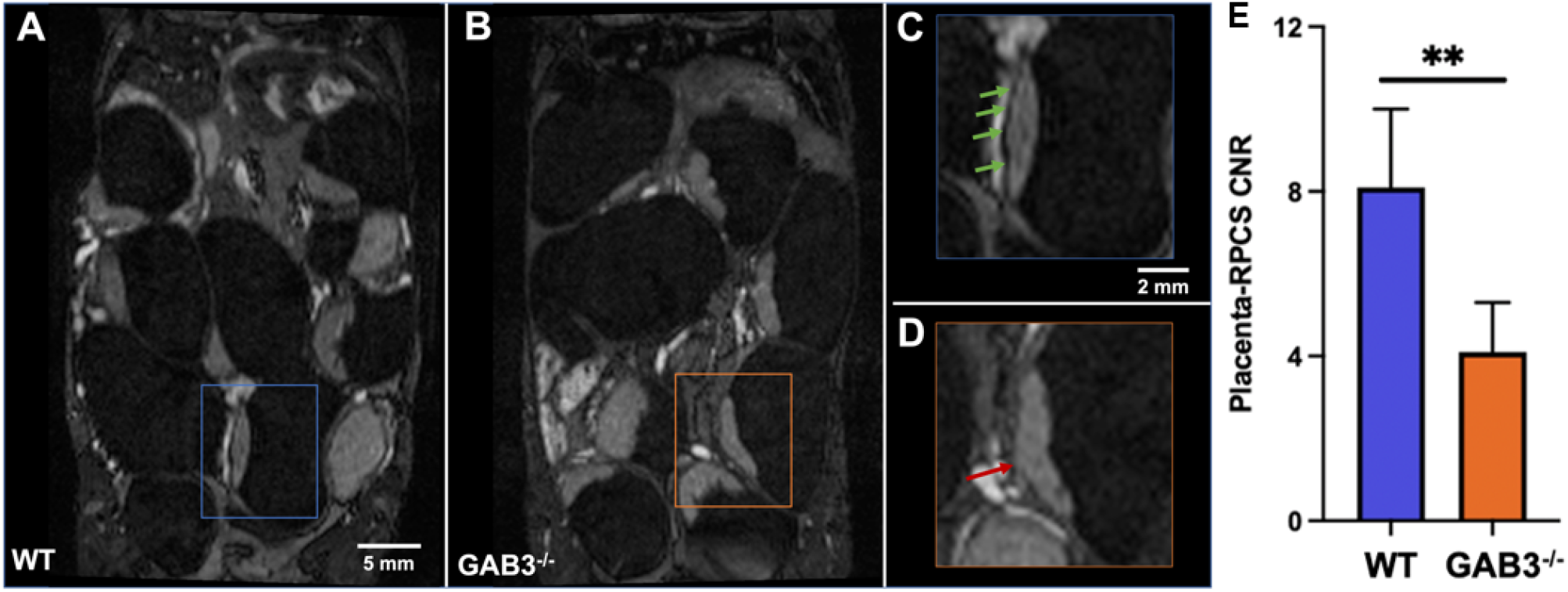
Ferumoxytol-enhanced MRI (Fe-MRI) of Gab3^-/-^ pregnant mice showed a diminished and heterogeneous retroplacental clear space (RPCS) when compared with control wild type (WT) pregnant mice. **(A)** Fe-MRI of WT mice shows enhancement in the mouse placenta, and **(C)** a hypointense region (green arrows) between placenta and myometrium that represents the RPCS. **(B)** Gab3^-/-^ mice also demonstrated high signal in placenta, however **(D)** the RPCS in Gab3^-/-^ pregnant mice appeared more jagged, thinner, and exhibited higher signal enhancement. **(E)** The CNR between placenta and RPCS is significantly lower in FPUs of Gab3^-/-^ mice than WT mice due to loss of RPCS and increased vascularity.

Ferumoxytol-enhanced MRI studies were conducted in patients considered at risk for PAS, and who required i.v. iron supplementation. In a severe PAS case with accreta (FIGO Grade 3C), initial Doppler ultrasound found significant hypervascularized structures, which were visible with non-contrast T2-weighted imaging (**Figure 4**). Fe-MRI images of the FIGO Grade 3C and a non-PAS (No FIGO score) case found frank areas of RPCS disruption and lower Placental-RPCS CNR and RPCS continuity volume (**Figure 4**). At term, the FIGO Grade 3C case demonstrated high degrees of utero-placental adherence and hypervascularity. In a second, moderate PAS case (FIGO Grade 1), ultrasound findings were unclear as were conventional T1 and T2-weighted non-contrast MR images (**Figure 5**). Fe-MRI, however detected disruption of the posterior portion of the RPCS along with areas of strong adhesion to the cervix. In this case, adherence at the cervical interface necessitated a hysterectomy at term.

**Figure 4:**
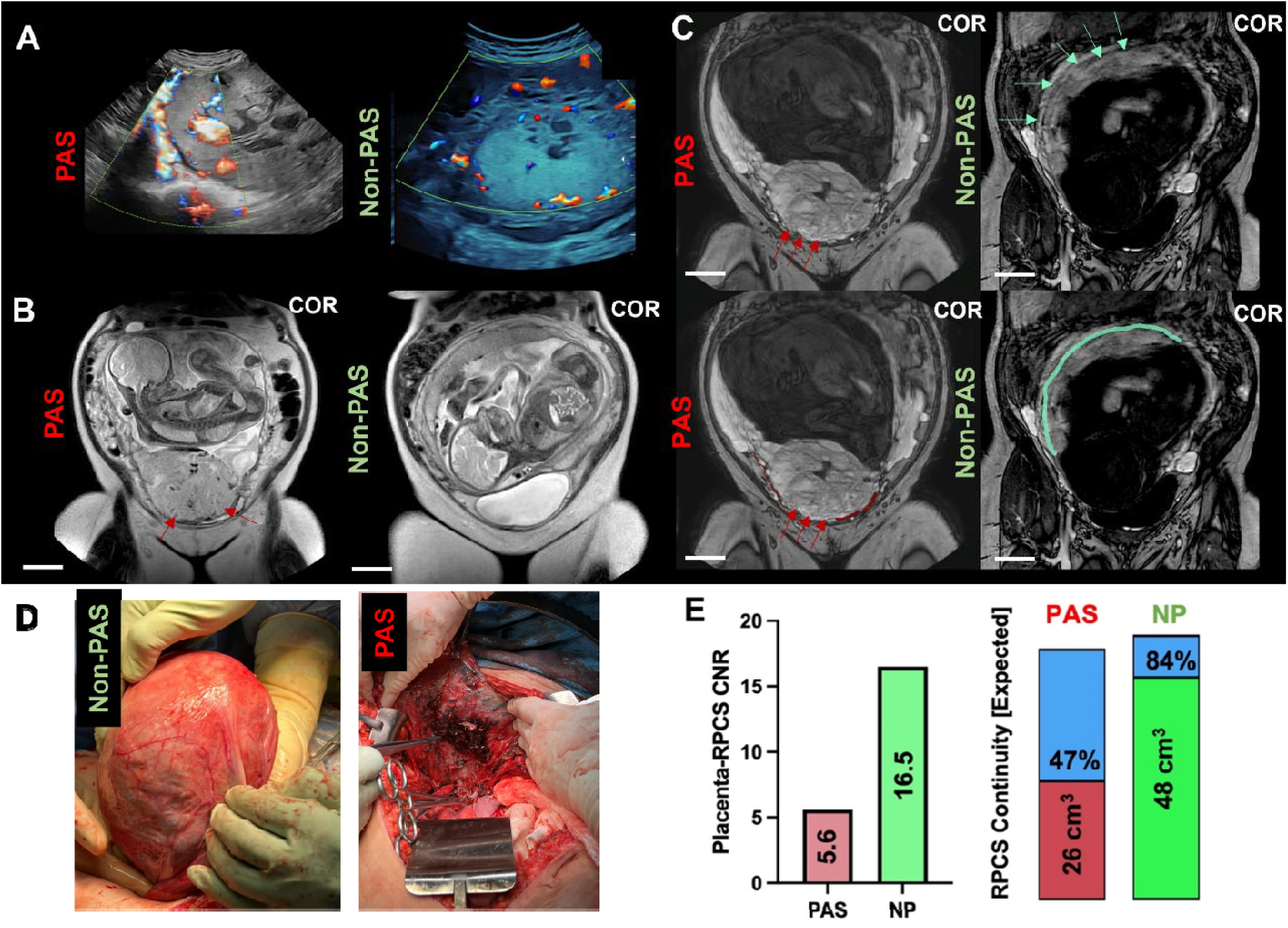
Fe-MRI enables visualization of severe PAS pathology in humans. (**A** Doppler ultrasound shows a greater degree of hypervascular structures in a PAS case relative to a non-PAS patient (Normal). (**B)** Coronal T2-weighted non-contrast MRI shows dark intraplacental T2 bands and frank invasion by the placenta. (**C)** Coronal T1-weighted mDixon images (post-contrast) enable delineation of the RPCS (green arrows and outlined) for both cases along with infiltration (red arrows). (**D)** Post-operative photos demonstrate differences in utero-placental adherence and vascularity. (**E)** Quantitative metrics include placental to RPCS contrast to noise ratio (CNR), which calculates the difference in signal-to-noise ratio for both regions, and comparison of RPCS continuity, where the color bar length is scaled by the expected volume for a non-interrupted RPCS path for both subjects. Scale bars denote 5 cm.

**Figure 5:**
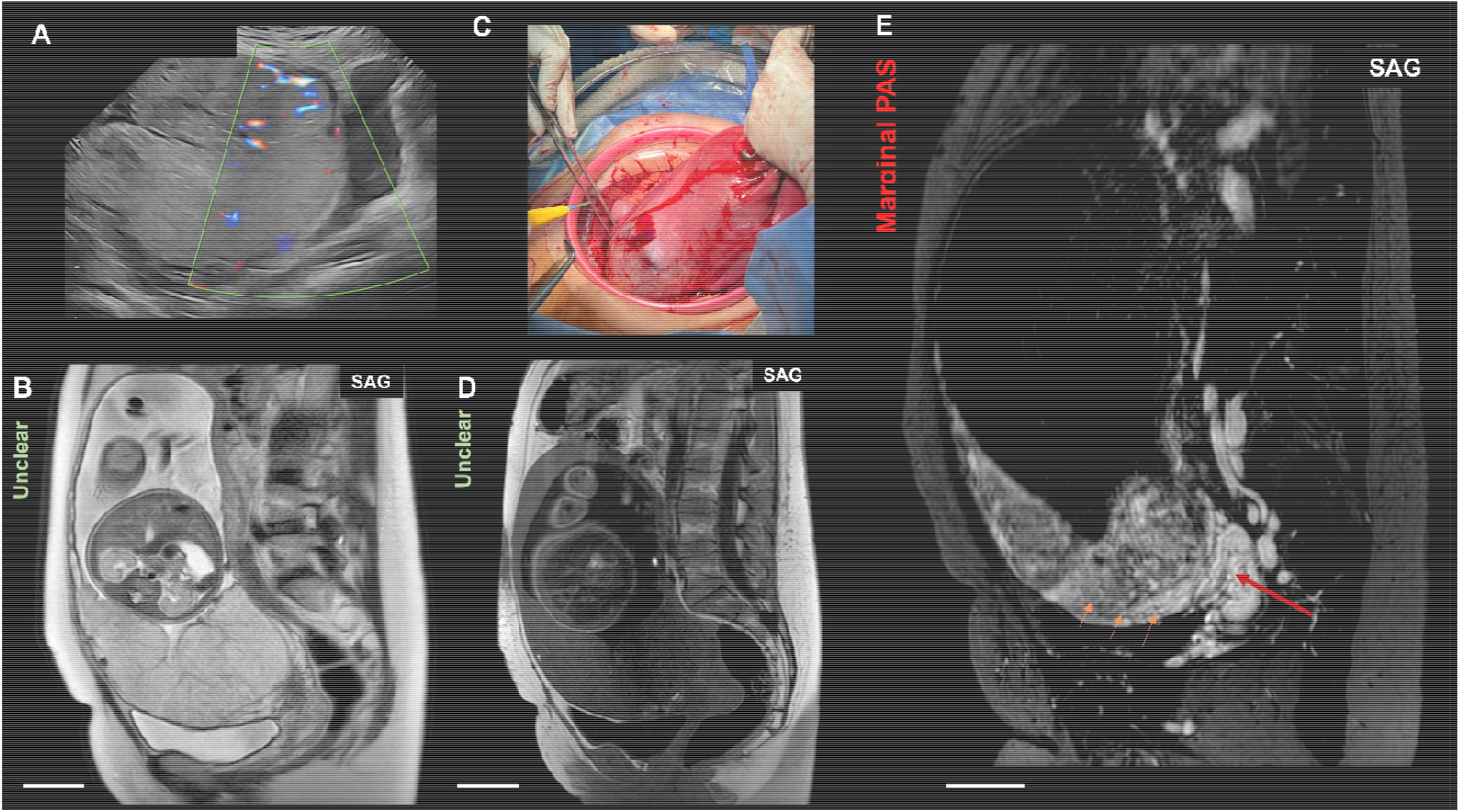
Visualization of a marginal PAS patient. **(A)** Despite the presence of previa diagnosed with ultrasound, (**B**) non-contrast T2-weighted imaging shows no dark intraplacental bands and no obvious invasion. (**C**) At term, however, strong invasion was seen near the cervical boundary and mandated a hysterectomy. **(D)** Non-contrast T1-weighted images did not show the level of scarring or adhesion. **(E)** Ferumoxytol-enhanced magnetic resonance angiography shows frank disruption of the RPCS (orange arrows) and areas of invasion/infiltration near the cervix (red arrow). Scale bars denote 5 cm.

In addition to visualizing the RPCS, Fe-MRI enabled high-resolution visualization of complete uteroplacental vasculature, which can enable assessment of both hyper- and hypo-vascularized regions of the placenta. In the non-PAS case, the superior portion of the placenta had avascular regions that were visible in ferumoxytol-enhanced images, but not conventional, non-contrast MRI (**Figure 6**). In the FIGO Grade 3C case, large, abnormal, tortuous vessels were visible on the surface of the placenta with Fe-MRI, and matched findings at term.

**Figure 6:**
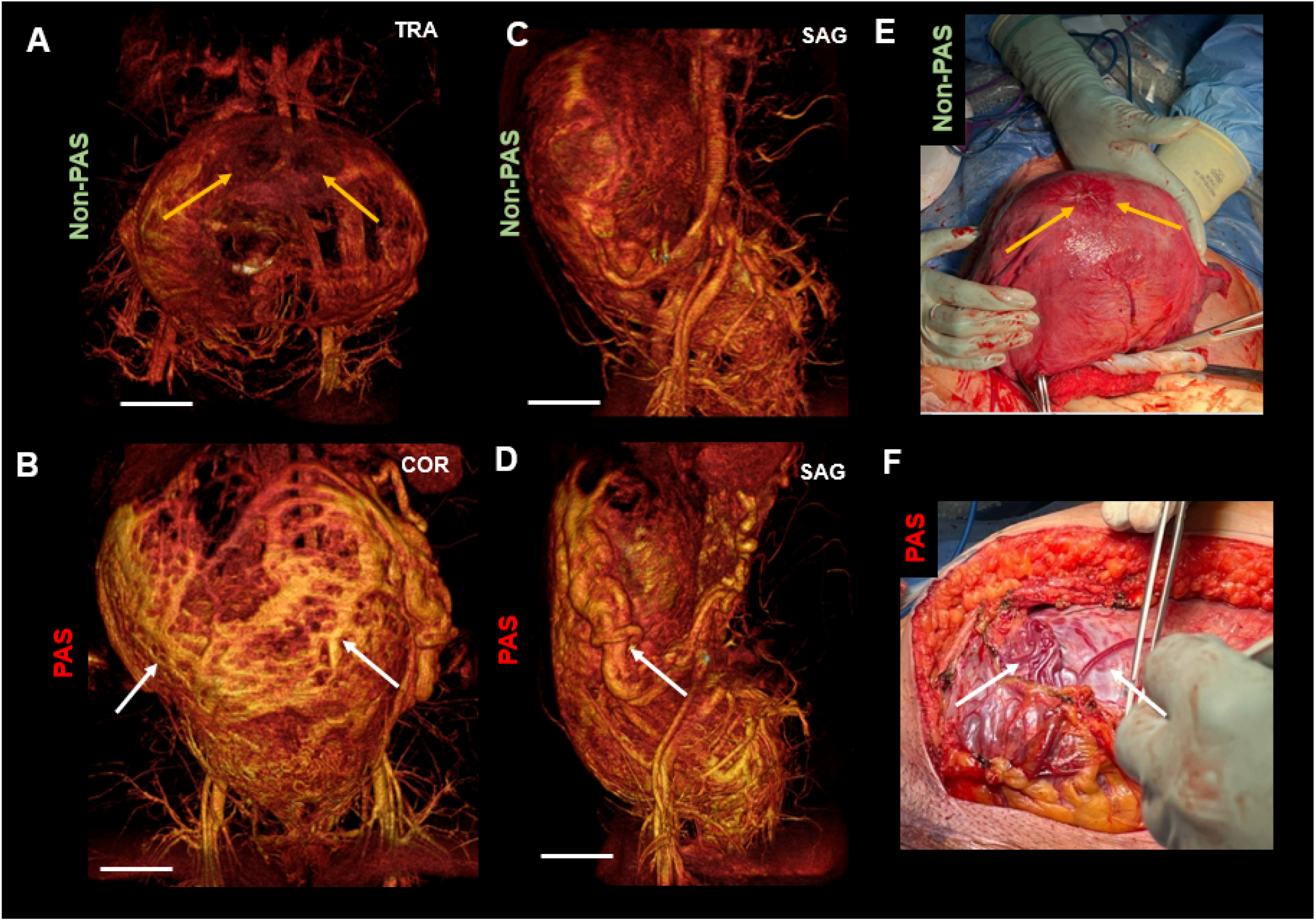
Fe-MRI enables visualization and quantification of uteroplacental hyper- or hypo-vascularity. **(A, C)** Volume renderings of 3D Fe-MRI data from a non-PAS patient show smaller caliber blood vessels that have less enhancement than a (**B, D**) PAS patient. Regions of hypovascularity are also visible in the non-PAS images (orange arrows). (**E-F**) Post-surgical images for both subjects demonstrate hypo- and hyper-vascularized nature of placenta in both cases.

## Discussion & Conclusions

Presented in this manuscript is a feasibility study demonstrating the potential of Fe-MRI to aid in diagnosis of PAS and presence of abnormal uteroplacental vasculature. Preclinical studies demonstrated that a lower dose of ferumoxytol (5 mg/kg) than normally used for iron supplementation (∼10-20 mg/kg) provides strong T1 shortening at clinically relevant MR field strength [28]. Fe-MRI enabled clear demarcation of the placental margins and increased SNR highlighting the hypointense RPCS, or uteroplacental interface (**Figure 2**). The Placenta-RPCS CNR, a ratio of the SNRs in the RPCS and placenta respectively, provides a quantitative measure to test the hypothesis that an interrupted and infiltrated RPCS exhibits greater T1w-MRI signal heterogeneity in Fe-MRI than one lacking obstruction, consistent with heterogeneous vascularization of the RPCS resulting from invasion of the placenta into the myometrial wall. The CNR measure yielded significant differences between WT and Gab3^-/-^ mice, similar to our previous work using liposomal-Gd contrast agent [15].

In patients, Fe-MRI demonstrated high intraplacental signal compared to conventional, non-contrast scans (**Figure 5D, Figure 5E**). In the FIGO Grade 3C case shown in Figure 4, conventional ultrasound detected an abnormally high degree of vascularity, and dark intraplacental bands were seen in conventional T2-weighted imaging. Fe-MRI scans showed multiple areas where the RPCS was completely obscured and discontinuous. In contrast, in the FIGO Grade 1 case that had unclear ultrasound and conventional non-contrast MRI findings, Fe-MRI scans showed presence of tortuous vessels and frank invasion close to the cervix, which were confirmed at delivery.

Clinically, PAS is correlated with placenta previa particularly in patients who previously underwent a cesarean section [29–32]. Yet, these cases are often not diagnosed by ultrasound and conventional non-contrast MRI, leading to uncertainty in surgical planning. A method that is capable of visualizing borderline cases of PAS in the setting of placenta previa would be exceptionally valuable for increasing the specificity of PAS diagnosis. Further, the ability to diagnose PAS in patients without placenta previa would provide tremendous benefit as these cases have a high probability of not being diagnosed prior to delivery [33]. Studies with Fe-MRI can provide such benefit, particularly since abnormal vascular development has been theorized as a potential predictor for maternal outcomes in accreta [34,35]. As shown in Figure 6, the FIGO 3C case has two important observations on Fe-MRI: 1) a high degree of placental signal relative to the non-PAS case, and 2) the presence of multiple large and tortuous vessels on the uteroplacental surface. Quantitative assessment of these features could provide an index for better differentiating the severity of PAS cases.

A central limitation of this study is the limited number of clinical datasets. Future studies will consider greater numbers of subjects across the range of PAS disorders and enable stronger conclusions grounded in quantitative metrics.

This study demonstrates the potential of ferumoxytol, an FDA-approved iron supplement, to provide strong T1-weighted contrast-enhanced MR images in pregnant patients. A contrast-enhanced MR technique with ferumoxytol could improve sensitivity for assessing the status of RPCS and increase the accuracy for diagnosis of PAS. Further, use of this technique will enable investigation into other mechanistic hypotheses surrounding abnormal placental development that may hold clues for better mitigation and interventional strategies.

## Data Availability

All data produced in the present study are available upon reasonable request to the authors

## Acknowledgments

The authors gratefully acknowledge the Cincinnati Children’s Hospital Medical Center and Dr. Helen Jones for providing the Gab3^-/-^ mouse model. The authors also acknowledge the contributions of MRI technologist Sarah Lawler and the Texas Children’s Hospital Radiology department. Financial support for the non-clinical portion of this study was provided by the National Institutes of Health (NIH) Grant R01HD094347.

